# *Prebiotic Inulin for Late-Reproductive Individuals (P.I.L.I. study):* A Protocol for a Placebo-Blind, Cross-Over Randomized Controlled Trial

**DOI:** 10.64898/2026.11.02.26367110

**Authors:** Mahya Mehri Hajmir, Stevaughn Smith, Mimi Ghosh, Philip Appiah, Adam Ciarglelio, Ali Rahnavard, Carmen P. Ortega-Santos

**Affiliations:** Exercise and Nutrition Sciences Department, Milken Institute School of Public Health, The George Washington University, Washington, DC, USA; Department of Epidemiology, George Washington University Milken Institute School of Public Health, The George Washington University, Washington, DC, USA; Department of Biostatistics and Bioinformatics, Milken Institute School of Public Health, George Washington University, Washington, DC, USA; Computational Biology Institute, Department of Biostatistics and Bioinformatics, Milken Institute School of Public Health, The George Washington University, Washington, DC, USA

**Author notes:** Correspondence concerning this article should be addressed to Carmen Ortega-Santos, Department of Exercise and Nutrition Sciences, Milken Institute School of Public Health, 950 New Hampshire Avenue, Office 211, Washington, DC 20037. These authors contributed equally to this manuscript (Mahya Mehri Hajmir, Stevaughn Smith).

**Keywords:** Gut Microbiome, Metabolome, Fiber, Women, Prebiotics, Clinical Trial Protocol, Inulin, Perimenopause, Healthy Aging

## Abstract

**Background:** Perimenopause is a transitional stage preceding menopause, characterized by hormonal fluctuations and physiological changes that may increase cardiometabolic risk. Emerging evidence suggests that the gut microbiome may influence these changes through estrogen metabolism and microbial metabolite production. Inulin, a prebiotic fiber that modulates the gut microbiome, may offer a targeted nutritional strategy; however, its effects during perimenopause remain largely unexplored.

**Objective:** This proof-of-concept study evaluates the effects of short-term inulin supplementation on gut microbiome composition, microbial metabolites, vascular function, skeletal muscle health, and metabolic flexibility in perimenopausal overweight women or with obesity.

**Methods:** We will conduct a randomized, blinded, placebo-controlled crossover trial among 27 women aged 40–55 years. Participants will receive 10 g/day of inulin and a matched maltodextrin placebo for two 14-day intervention periods separated by a 2–4-week washout. We will assess changes in gut and vaginal microbiome composition and functional capacity, stool and plasma microbial metabolites, and circulating markers of estrogen metabolism as primary outcomes. We will also evaluate vascular function, skeletal muscle health and oxidative capacity, and metabolic flexibility as secondary outcomes. Additionally, we will assess physical activity, sleep, dietary intake, perimenopausal symptoms, and intervention feasibility, adherence, acceptability, and tolerability.

**Conclusion:** This study will provide novel insights into the feasibility and biological effects of inulin supplementation on gut-host mechanisms and cardiometabolic health during perimenopause, informing the design of future larger-scale trials.

## Introduction

The menopausal transition is a life stage that all women will experience, with unique implications for hormonal, physiological, and metabolic health that can significantly accelerate the risk of cardiometabolic diseases. [1–3] Physiological hallmarks of the menopausal transition include increased insulin resistance, loss of lean mass, vascular dysfunction, and declining vaginal health.[4–8] Emerging research highlights the gut *estrobolome,* i.e., gut-synthesized bioactive fecal and circulating metabolites related to estrogen, as a novel factor contributing to sex hormone imbalances and as a potential significant contributor to the increase in cardiometabolic risk factors in the menopausal transition.[9–13] Because diet is the primary factor shaping gut microbial composition and the resulting metabolome,[14] this presents an opportunity to develop targeted nutritional interventions to improve perimenopausal health and support healthy aging.

Inulin is a prebiotic fiber used as a nutraceutical for its beneficial effects on cardiometabolic risk factors in premenopausal women with obesity and altered estrogen metabolism, mediated by the gut microbiome.[15,16] Previous studies have demonstrated that inulin supplementation alters circulating and fecal metabolite profiles, including polyamines, amino acid metabolites, short-chain fatty acids, bile acid metabolites, and lipid-derived compounds, with several of these changes associated with shifts in metabolic health outcomes.[17,18] However, the metabolomic response to inulin supplementation during the perimenopausal transition remains largely unexplored. This study will evaluate, as a proof of concept, the effects of inulin on the gut estrobolome and gut microbial composition among overweight women or women with obesity.

In addition to the changes in the gut microbiome that influence cardiometabolic health during perimenopause, this life stage is also characterized by declines in skeletal muscle function, impaired metabolic flexibility, and worsening vascular health.[19,20] These physiological systems are closely linked to estrogen status and may also be influenced by metabolites derived from the gut microbiome. Therefore, alongside primary gut microbiome outcomes, this study will explore inulin’s effects on muscle function, metabolic flexibility, and vascular health to provide a more comprehensive evaluation of its potential benefits during the menopausal transition.

The current literature increasingly recognizes perimenopause as a critical period for musculoskeletal decline in women. Emerging evidence suggests that symptoms, including hormonal changes, may be linked to reductions in muscle mass, strength, and neuromuscular function. A large systematic review and meta-analysis of over 93,000 women found that musculoskeletal symptoms, including muscle and joint pain, are common but often underreported across the stages of perimenopause. Consistent with this, a study of over 400 peri- and postmenopausal women reported that lower muscle mass was associated with more severe menopausal symptoms, an effect partially mediated by estrogen levels.[21] Importance of muscle health as a key, historically overlooked, dimension of women’s health during perimenopause.

As estrogen levels gradually fall during perimenopause, there is an associated disruption in metabolic flexibility, referring to the body’s adaptive ability to shift from fatty acid oxidation to glucose oxidation in response to metabolic demand.[22] Because skeletal muscle is the primary site of substrate metabolism, decreases in muscle mass or quality triggered by the menopausal transition can impair metabolic processes and increase the risk of insulin resistance and other metabolic diseases, such as diabetes, especially in this population, along with its associated undesired alterations to body composition and fat distribution patterns.[23]

Similarly, vascular health is negatively affected by the menopause transition. Physiologically, estrogen supports endothelial function and vascular elasticity. So, as estrogen levels gradually decrease and ovarian function declines during the menopause transition, vascular aging accelerates, which then raises risk factors such as endothelial dysfunction and increased arterial stiffness, contributing to cardiovascular disease in women.[24] Based on the literature, this research study evaluates the effects of inulin prebiotic fiber on the gut and vaginal microbiome and estrobolome to ameliorate cardiovascular and metabolic symptoms during the menopausal transition among women with overweight and obesity.

### Aims

#### Primary Objective

- The first primary objective is to determine whether a 2-week inulin supplementation period, compared with a matched maltodextrin placebo, alters gut and vaginal microbiome composition and functional capacity in perimenopausal women who are overweight or women with obesity. We hypothesize that inulin supplementation will improve microbiome composition and function, including estrobolome-related microbial pathways and metabolites, compared with placebo.
- The second primary objective is to determine whether a 2-week inulin supplementation period, compared with placebo, affects microbial metabolite profiles and circulating markers of estrogen metabolism. We hypothesize that inulin supplementation will result in favorable changes in stool and plasma microbial metabolites and estrogen-related biomarkers compared with placebo.

#### Secondary Objective

- The first secondary objective is to determine whether a 2-week inulin supplementation period, compared with placebo, improves vascular function in perimenopausal women with overweight or obesity. We hypothesize that inulin supplementation will improve vascular function, assessed by pulse-wave velocity and resting heart rate, compared with placebo.
- The second secondary objective is to determine whether a 2-week inulin supplementation period, compared with placebo, improves skeletal muscle health in perimenopausal women who are overweight or women with obesity. We hypothesize that inulin supplementation will improve skeletal muscle oxidative capacity, as assessed by near-infrared spectroscopy, and muscle function, as assessed by handgrip strength, compared with placebo.
- The third secondary objective is to determine whether a 2-week inulin supplementation period, compared with placebo, improves metabolic flexibility in perimenopausal women with overweight or obesity. We hypothesize that inulin supplementation will improve metabolic flexibility, assessed during a mixed-meal challenge, compared with placebo.

#### Exploratory Objectives

The exploratory objectives of this study are to characterize the effects of a 2-week inulin supplementation period on lifestyle behaviors and perimenopausal symptom trajectories. Exploratory outcomes will include objectively measured physical activity and sleep characteristics assessed using Fitbit Inspire 3-derived metrics, subjective sleep quality assessed using the Pittsburgh Sleep Quality Index, and weekly changes in perimenopausal symptoms.

#### Feasibility Objectives

- The feasibility objectives of this study are to evaluate the feasibility, acceptability, adherence, and tolerability of the intervention and study procedures to inform the design of a future adequately powered randomized controlled trial.

## Methods

### Study Design

The PILI study is a proof-of-concept, randomized, blinded, placebo-controlled, cross-over trial designed to evaluate the effects of inulin supplementation on the gut microbiome, gut-derived metabolites, skeletal muscle mitochondrial oxidative capacity, and cardiometabolic health in perimenopausal women with overweight or obesity. Participants will be randomly assigned to one of two intervention sequences (inulin followed by placebo [AB] or placebo followed by inulin [BA]). Each intervention period will last two weeks and be separated by a two-to four-week washout period to minimize potential carryover effects. Because each participant receives both interventions, each serves as her own control, reducing between-subject variability and improving the ability to detect within-subject changes.

Outcome assessments will be conducted before and after each intervention period using standardized study procedures. Participants, investigators, and study personnel involved in outcome assessment will remain blinded to treatment allocation throughout the study. In addition to evaluating preliminary biological effects of inulin supplementation, the study will assess the feasibility, acceptability, adherence, and tolerability of the intervention and study procedures to inform the design of a future adequately powered randomized clinical trial.

### Study Setting

This study will be implemented in the Washington, DC metropolitan area through the Milken Institute School of Public Health at George Washington University. Study activities, including participant recruitment, intervention delivery, health assessments, biological sample collection, and data management procedures, will be conducted at approved community and university research sites participating in the project. Laboratory processing and analytical procedures will be completed at the university and within university-affiliated research facilities.

### Participants

The study will enroll perimenopausal overweight women or with obesity who meet the eligibility criteria described below. All participants will provide written informed consent before completing any study procedures.

*Eligibility criteria.* Table 1 lists the inclusion and exclusion criteria.

**Table 1.** Participants inclusion and exclusion criteria.

| **Inclusion Criteria** |
|---|
| Biological Sex: Women |
| 40 – 55 years old |
| Body mass index: 25 – 40 kg/m<sup>2</sup> |
| Menstrual cycle irregularity (cycle length outside 21 – 35 days or variation > 7 days) or/and 2- 11 months of amenorrhea; with or without hot flashes or/and night sweats |
| Weight stable within the last 3 months (body weight fluctuations within 5% of their habitual body weight) |
| Spending ≥ 6 hours sitting |
| < 150 minutes per week (50 minutes per day, 3 days per week OR 30 minutes per day, 5 days per week) of moderate to vigorous structured physical activity (i.e., planned intentional physical activity, not incidental movement). |
| **Exclusion Criteria** |
| Recent antibiotic use oral or vaginal (≥ 6months); |
| Diagnosed with any chronic disease (e.g., any gastrointestinal condition like IBD, type 2 diabetes, or cancer) |
| Prescription of hormone replacement therapy, hormonal contraceptives, probiotics, GLP-1 medications, or weight loss medications |
| Smoking |
| Recent blood donation (within the past 8–12 weeks) |
| Significant weight changes (≥5%) in the last 4 weeks |
| Use of dietary supplements (unless discontinued 4 weeks prior) |
| Participants taking long-term, stable medications for chronic conditions such as hypertension or hyperlipidemia will *not* be excluded if their medication regimen has been unchanged for at least 2 years, as these treatments are part of their usual health status. Recent medication changes (within the past 6 months), initiation of new prescriptions, or use of medications known to affect metabolic, vascular, vaginal, hormonal, or gastrointestinal function will result in exclusion. |
| Fructose intolerance |

### Recruitment

We will recruit participants from the Washington, DC area using community-based and university strategies, including flyers, email lists, social media, community groups, and university channels. We will use the ResearchMatch platform for targeted research recruitment. Interested individuals will complete an online eligibility screening. The study will target community women aged 40–55 with overweight or obesity, and progress will be monitored to recruit 27 participants, aiming for 20 completers.

### Randomization and Blinding

Participants will be randomly assigned in a 1:1 ratio to one of two intervention sequences: Sequence AB (inulin followed by placebo) or Sequence BA (placebo followed by inulin). The study statistician generated the sequence allocation schedule before trial initiation using a predetermined, computer-generated randomization table.

This study employs a participant- and analyst-blinded protocol. Participants remain blinded to treatment sequence throughout the trial, while study investigators responsible for supplement management and participant visits are unblinded. To maintain participant blinding, both inulin and maltodextrin placebo are packaged in identical containers with matching appearance, weight, dosing instructions, and administration schedules.

### Intervention

Participants will receive either inulin supplementation (Sensus Inulin, Sensus America Inc., NJ) or a matched maltodextrin placebo for two 14-day intervention periods in a randomized crossover design. Inulin is a fermentable dietary fiber that serves as a substrate for gut microbial fermentation and may influence microbiome composition, microbial functional capacity, and metabolite production.^20^

For each intervention period, participants will receive one bottle containing 80 g of the assigned supplement, along with a food scale and measuring scoop to facilitate accurate dosing.

Participants will be instructed to weigh and consume 10 g of the assigned supplement daily for 14 consecutive days and to track the amounts in their electronic diary. Any remaining supplement will be returned in the original container at the end of each intervention period to assess adherence by comparing the dispensed and returned product weights.

Participants will be instructed to maintain their usual dietary intake and physical activity patterns throughout the intervention periods. Study staff will monitor adherence through supplement logs and the weight of returned supplements. Participants will also complete daily and weekly questionnaires to monitor gastrointestinal symptoms, tolerability, and intervention acceptability. Study staff will conduct weekly 24-hour dietary recalls, and participants will complete a perimenopause symptoms questionnaire.

### Study Procedures and Assessments

#### Screening

Potential participants will complete an online screening questionnaire to assess eligibility. Individuals meeting all inclusion criteria and none of the exclusion criteria will automatically receive an email to complete the short version of the International Physical Activity Questionnaire (IPAQ)[25] and a short medical history questionnaire via Research Electronic Data Capture (REDCap).[26] Upon confirmation of eligibility, participants will receive an email containing a copy of the informed consent document and a link to schedule a Zoom meeting with study personnel. Participants will have the opportunity to review the consent form before the scheduled meeting. During the Zoom session, study staff will confirm eligibility, review study procedures, answer participants’ questions, and obtain electronic informed consent from individuals who agree to participate.

#### Study Procedures

Participants will complete the same assessment battery before and after each intervention period. The assessment battery will consist of two in-person visits separated by 48–72 hours: (1) a vascular and musculoskeletal assessment visit and (2) a metabolic flexibility assessment visit. We will repeat this two-visit assessment battery under identical standardized conditions (including an overnight fast [≥ 8 hours], standardized pre-visit instructions, and assessment at approximately the same time of day whenever feasible) before and after both intervention periods. Unless otherwise specified, we will repeat the procedures described below at the corresponding pre- and post-intervention visits during both intervention periods. **Table 2** describes the study assessments and procedures.

**Table 2.**
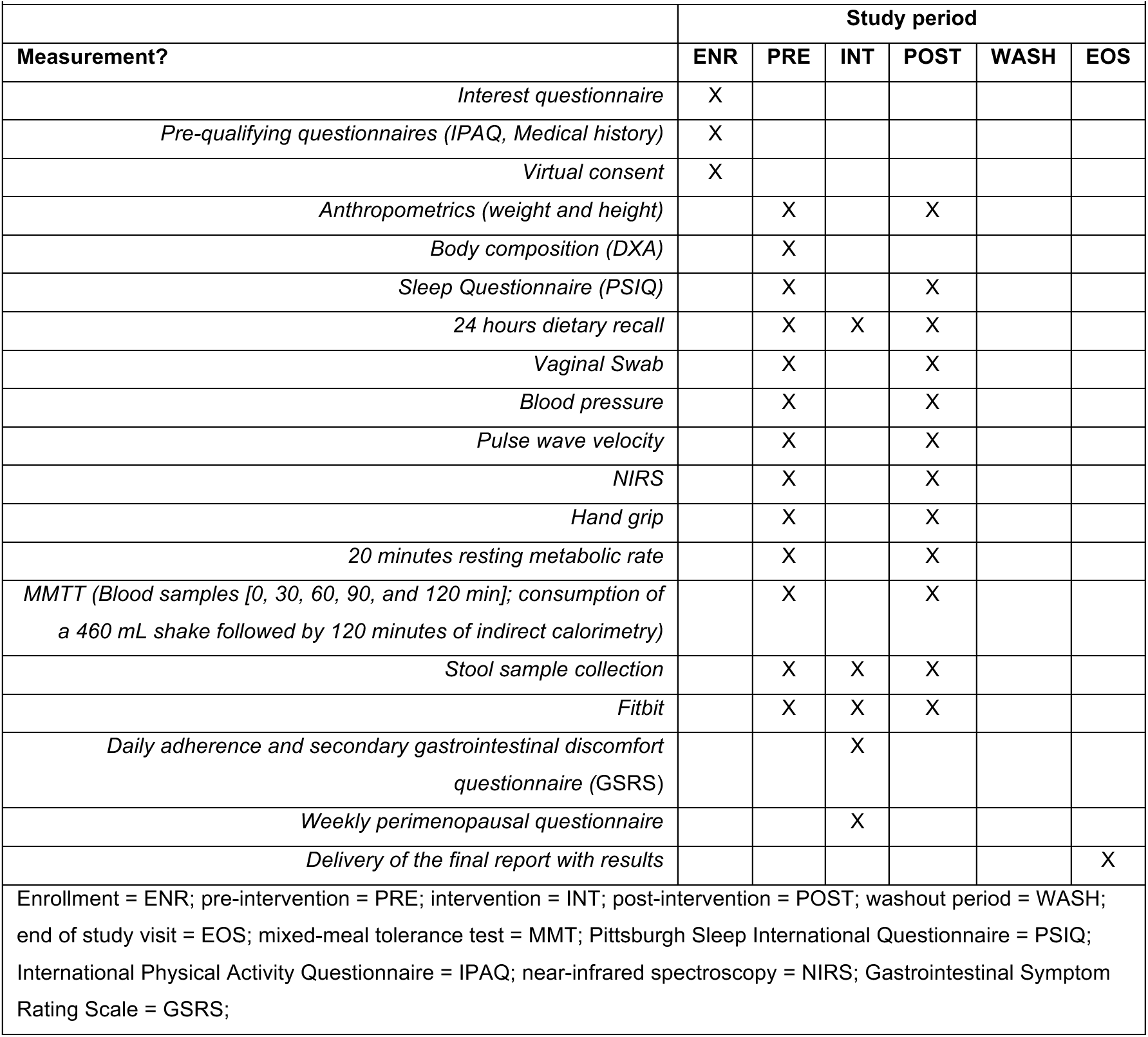
Schedule of enrollment, interventions, and assessments.

|  | Study period |  |  |  |  |  |
|---|---|---|---|---|---|---|
| **Measurement?** | **ENR** | **PRE** | **INT** | **POST** | **WASH** | **EOS** |
| *Interest questionnaire* | X |  |  |  |  |  |
| *Pre-qualifying questionnaires (IPAQ, Medical history)* | X |  |  |  |  |  |
| *Virtual consent* | X |  |  |  |  |  |
| *Anthropometrics (weight and height)* |  | X |  | X |  |  |
| *Body composition (DXA)* |  | X |  |  |  |  |
| *Sleep Questionnaire (PSIQ)* |  | X |  | X |  |  |
| *24 hours dietary recall* |  | X | X | X |  |  |
| *Vaginal Swab* |  | X |  | X |  |  |
| *Blood pressure* |  | X |  | X |  |  |
| *Pulse wave velocity* |  | X |  | X |  |  |
| *NIRS* |  | X |  | X |  |  |
| *Hand grip* |  | X |  | X |  |  |
| *20 minutes resting metabolic rate* |  | X |  | X |  |  |
| *MMTT (Blood samples [0, 30, 60, 90, and 120 min]; consumption of a 460 mL shake followed by 120 minutes of indirect calorimetry)* |  | X |  | X |  |  |
| *Stool sample collection* |  | X | X | X |  |  |
| *Fitbit* |  | X | X | X |  |  |
| *Daily adherence and secondary gastrointestinal discomfort questionnaire (GSRS)* |  |  | X |  |  |  |
| *Weekly perimenopausal questionnaire* |  |  | X |  |  |  |
| *Delivery of the final report with results* |  |  |  |  |  | X |

Enrollment = ENR; pre-intervention = PRE; intervention = INT; post-intervention = POST; washout period = WASH; end of study visit = EOS; mixed-meal tolerance test = MMT; Pittsburgh Sleep International Questionnaire = PSIQ; International Physical Activity Questionnaire = IPAQ; near-infrared spectroscopy = NIRS; Gastrointestinal Symptom Rating Scale = GSRS;

### Outcomes

#### Primary outcomes

Change in gut microbiome composition and functional capacity, including biosynthetic gene clusters (BGCs)[27] and microbial pathways involved in estrogen metabolism (estrobolome), vaginal microbiome composition, and stool and plasma microbial metabolites following 2 weeks of inulin supplementation compared with placebo. We will characterize gut and vaginal microbiome profiles using shotgun metagenomic sequencing and quantify microbial metabolites using targeted metabolomic analyses.

#### Secondary outcomes

Secondary outcomes will evaluate the effects of inulin supplementation on skeletal muscle health, cardiometabolic health, lifestyle behaviors, and perimenopausal symptoms. We will assess skeletal muscle health using handgrip strength and mitochondrial oxidative capacity in skeletal muscle, measured with near-infrared spectroscopy (NIRS).[28,29] We will evaluate cardiometabolic health through vascular function (pulse wave velocity [PWV] and resting heart rate) and metabolic flexibility (assessed during a mixed-meal challenge). Lifestyle behaviors will include objectively measured physical activity using a Fitbit accelerometer and sleep quality, assessed using the Pittsburgh Sleep Quality Index (PSQI)[30] and the Fitbit-derived sleep parameters like the sleep metric score, total sleep duration, sleep efficiency, and time spent in sleep stages (light, deep, and rapid eye movement [REM] sleep).[31] Perimenopausal symptom severity will be assessed weekly using a recently developed 16-item perimenopause symptom questionnaire.[32]

#### Feasibility outcomes

The primary feasibility outcomes will include recruitment, retention, adherence, acceptability, and tolerability of the intervention and study procedures. Recruitment feasibility will be evaluated by the recruitment rate (participants enrolled per month), eligibility rate (eligible participants among those screened), enrollment rate (participants enrolled among those eligible), and reasons for ineligibility or declining participation. Retention will be assessed by study completion and attrition rates, as well as reasons for participant withdrawal.

Intervention adherence will be evaluated through adherence to the assigned inulin or placebo supplementation, assessed by returned bottle counts and participant daily logs, as well as adherence to scheduled study visits, home stool sample collection, and completion of daily and weekly study questionnaires. Acceptability and tolerability will be assessed throughout the intervention by monitoring daily gastrointestinal symptoms using an adapted GSRS 15-item questionnaire, including bloating, gas, abdominal discomfort, and other self-reported gastrointestinal complaints.[33] Weekly questionnaires will evaluate participants’ perceived acceptability and tolerability of the intervention and study procedures. These feasibility outcomes will be used to assess the practicality of the study protocol and inform the design of a future adequately powered randomized controlled trial.

### Data analysis

#### Clinical and Physiological Outcomes Analysis

Blood samples collected pre-and post-intervention at the baseline blood draw during the MMTT will be used to analyze a complete panel of sex hormones, including follicle-stimulating hormone, luteinizing hormone, estrogen, progesterone, and testosterone, using commercially available ELISA kits. The samples will be processed, plasma separated by centrifugation, and stored at -80°C for later analysis.

#### Metagenomics Analysis

Vaginal and stool samples will undergo metagenomic analysis. Initial processing involves extracting genomic DNA from the collected stool samples using the ZymoBIOMICS DNA Miniprep Kit (Zymo, CA, US). Then, DNA will be sequenced by shotgun metagenomics with the NextSeq 2000 High-Output 300-cycle kits (Illumina), followed by standardized data filtering and control protocols.

#### Metabolomics Analysis

Whole blood will be immediately processed and centrifuged to obtain plasma. Plasma homogenates will undergo automated biochemical extraction to isolate the relevant metabolites. After extraction, we will analyze the metabolites using gas chromatography coupled with high-resolution tandem mass spectrometry (GC-MS/MS).

#### Sample Size Determination

Because this is a proof-of-concept pilot study, we did not perform a formal prospective sample size calculation. We selected a target sample size of 27 enrolled participants (aiming for at least 20 completers, accounting for an estimated ∼25% attrition rate) based on feasibility, participant burden, and published recommendations for pilot crossover trials evaluating multi-omics and physiological endpoints.[34] A primary objective of this trial is to obtain preliminary estimates of effect sizes (e.g., Cohen’s *d*, partial *n^2^*) and within-subject variability for gut microbiome composition, microbial metabolite concentrations, vascular function, and metabolic flexibility metrics. We will use the effect sizes and confidence intervals from this study to conduct formal power and sample size calculations for a subsequent, fully powered large-scale clinical trial.

#### Statistical Analysis

We will evaluate feasibility outcomes, including recruitment rates, eligibility rates, participant retention, supplement adherence, protocol adherence, acceptability, and gastrointestinal tolerability (GSRS scores), using descriptive statistics (frequencies, percentages, means with standard deviations, and medians with interquartile ranges). We will use a blinded crossover design with randomized treatment order, assigning participants to a pre-concealed sequence to ensure blinding. A 2-week wash-out minimizes carryover. This design enhances internal validity by increasing power and controlling confounding factors. We will summarize the data using descriptive statistics, test for normality using the Kolmogorov-Smirnov test and Q-Q plots and handle missing data through multiple imputations as needed. For outcomes measured across the four time points (baseline, post-intervention, post-washout, and post-placebo), we will use linear mixed-effects models, with a random intercept for participant ID to account for within-subject correlations. This approach will allow us to examine the impact of time and intervention (inulin vs. placebo) on the outcomes of interest while adjusting for baseline values and relevant covariates. We will use Permutational Multivariate Analysis of Variance to compare overall microbiome composition across time points and interventions. Differential abundance analyses using Multivariate Microbial Association with Linear Models will assess changes in specific bacterial taxa and biosynthetic gene clusters encoding GUS enzymes.[35] Due to the unique properties of multi-omics data (e.g., zero-inflation and high dimensionality), we will use machine-learning-based techniques, including: 1) *omeClust*, for identifying patterns in omics data; 2) *Tweedieverse*, for differential analysis to link omics features with outcomes like fitness level; 3) *omePath*: for pathway enrichment analysis and 4) *btest* tool to link, rank, and visualize associations among -omics features.

### Funding and Ethical Considerations

This study is funded by the intramural research grants Research Innovation Awards (05RIA112025) in the Milken Institute School of Public Health. The George Washington University Institutional Review Board (IRB) reviewed and approved this study protocol (NCR267325). All participants will provide written informed consent before any study procedures begin. The study will follow the ethical principles outlined in the Declaration of Helsinki and applicable institutional and federal regulations.

Participant confidentiality will be maintained through unique study identification numbers, and all study data and biospecimens will be stored with de-identified identifiers. Authorized study personnel will have access to identifiable information. The study team will monitor potential risks associated with the intervention and procedures throughout the study and will document and report adverse events in accordance with IRB requirements.

## Discussion

This proof-of-concept blinded RCT has limitations to consider when interpreting findings and designing future trials. The 2-week inulin period is brief and may not capture long-term microbiome or metabolic changes, but it will capture rapid compositional and functional changes that might indicate long-term directional shifts.[36] Longer interventions (over 26 weeks) are needed to assess the durability and significance of effects. The study lacks long-term follow-up, so it cannot evaluate sustained effects. Further, sex hormones in blood will be measured only pre- and post-intervention, therefore not capturing hormonal fluctuations that can occur during perimenopause. Future trials should use dried blood spot kits for frequent home sampling.

These limitations are inherent to the proof-of-concept design and should inform future trial planning, including extending duration to 4–12 weeks, adding long-term follow-up, controlling diet more rigorously, and capturing hormone fluctuations. Despite limitations, this study provides novel data using advanced multi-omics and offers feasibility insights for larger trials. Strengths include each participant serving as her own control to reduce variability; a 2–4-week washout to minimize carryover effects; blinding to prevent bias; and state-of-the-art techniques such as shotgun sequencing and metabolomics. Assessments under standardized conditions and feasibility metrics will inform future trial design. Enrolling overweight or obese perimenopausal women targets a high-risk group for metabolic interventions, relevant to public health. This proof-of-concept randomized, blinded, controlled, crossover trial will provide novel insights into the feasibility and biological effects of short-term inulin supplementation on the gut microbiome, microbial metabolites, vascular function, and metabolic flexibility in perimenopausal women with overweight or obesity. The menopausal transition represents a critical window characterized by increased susceptibility to weight gain, metabolic dysfunction, and cardiovascular risk [1,19]; however, effective and accessible strategies to mitigate these changes remain limited. By investigating the role of a targeted dietary fiber intervention on gut microbial function and cardiometabolic health, this study may advance our understanding of the gut–host mechanisms underlying midlife metabolic changes. Findings from this study will inform the design of future larger-scale clinical trials evaluating dietary approaches to support metabolic and vascular health in women during the menopausal transition.

## Supporting information

SPIRIT 2015 Checklist

## Author Contributions

**Conceptualization:** MG, CPOS. **Methodology:** MG, AC, AR, CPOS. **Formal analysis:** AC, AR, PA. **Investigation:** CPOS, MMH, SS. **Supervision & Project Administration:** CPOS. **Writing – original draft:** MMH, SS, MG, CPOS. **Writing – review & editing:** MG, PA, AC, AR, CPOS.

## Trial Registration

This study was registered on July 15^h,^ 2026, at clinicaltrials.gov.gov, ID: NCT07724522

## Data Availability

No data has been produced for this manuscript

